# Evaluating the Quality of Diabetes Care

**DOI:** 10.64898/2026.11.06.26367444

**Authors:** Prince Kenneth Homiah, Bernice Ameyaw, Kwamena Kwentsir Daniels, Ebenezer Mensah Turkson, Valerie Asare Baffour, Samuel Bekoe, Benjamin Oppong Nyantakyi

**Affiliations:** The Laboratory Unit, Ghana Ports and Harbours Authority Hospital, Takoradi, Ghana

**Keywords:** Diabetes, RTEC, albuminuria, Ghana

## Abstract

**Background:** Diabetes mellitus (DM) is an escalating global public health concern, contributing significantly to global mortalities and posing a major financial burden, particularly in Low- and Middle-Income Countries (LMICs) like Ghana [Letsa, 2023]. Suboptimal quality of care can lead to severe microvascular complications such as diabetic nephropathy (DN) and opportunistic infections like urinary tract infections (UTIs) [Letsa, 2023]. DN affects 30–40% of diabetics and is a leading cause of end-stage kidney failure, with a reported prevalence of kidney diseases of 12.5% in Ghana [Adoba et al., 2025]. Given that monitoring quality of care is crucial for prevention, this study aimed to evaluate the quality of DM care indicators, particularly focusing on urinary biomarkers, at the Ghana Ports and Harbours Authority (GPHA) hospital.

**Methodology:** A cross-sectional design employing a retrospective medical audit was utilized, assessing patient records at the GPHA hospital Diabetes Clinic. The study population comprised 125 participants whose urinalysis data, demographics, and comorbidity categorization (Diabetes only, Diabetes and Hypertension, Diabetes and Cyesis, etc.) were collected. Urine parameters, including albuminuria, glucose, and Renal Tubular Epithelial Cells (RTEC), were retrieved from Sysmex’s urinalysis analyzers (Sysmex UC 400, UF 350).

**Results:** The study cohort consisted predominantly of females (58.4%, n=73) with a mean age of 54.92 years. The primary diabetic categorization was “diabetes only” (76.8%, n=96). Evaluation of albuminuria showed that **mild albuminuria was recorded for only 10.4%** (n=13) of participants, and overt albuminuria was absent (0.0%). This mild albuminuria prevalence is notably below the 25–30% global prevalence reported for type 2 diabetics. A significant correlation was established between the various diabetes population categories and urine glucose (Pr=0.001). Pathological RTEC results (> 2.5) were identified in three individuals, primarily from the diabetes and hypertension category (2 individuals) and the diabetes and cyesis category (1 individual). The study noted limitations, including the difficulty in acquiring the specific type of diabetes and the non-availability of data on medication type or complication level.

**Conclusion:** The prevalence of mild albuminuria among the study population at the GPHA hospital was significantly lower than established global and certain regional Ghanaian rates. However, the detection of a significant association between diabetic categorization and glucosuria underscores the need for continuous and rigorous monitoring of renal function markers. These findings suggest that while indicators like mild albuminuria may be lower than expected in this specific cohort, targeted interventions remain essential to prevent the progression of renal complications in this high-risk population [ Adoba et al., 2025].

## Introduction

Diabetes mellitus (DM) is a rapidly escalating global health crisis, responsible for a significant increase in mortalities since the early 2000s and now ranking among the top ten causes of death worldwide. In 2021, the International Diabetes Federation (IDF) reported 537 million adults aged 20 to 79 living with diabetes, a figure projected to rise to 783 million by 2045, with 94% of this increase anticipated in low- and middle-income countries (LMICs) [Letsa, 2023]. Sub-Saharan Africa (SSA) is expected to experience the fastest rise in diabetes prevalence, facing a “double burden of disease” where diabetes exacerbates infection risks and strains limited healthcare resources [Letsa, 2023]. The global expenditure on diabetes reached 966 billion dollars in 2021, with Africa alone accounting for approximately 13 billion dollars [Letsa, 2023].

Diabetes is associated with severe macrovascular and microvascular complications [Letsa, 2023]. Diabetic nephropathy (DN), a major microvascular complication, affects 30–40% of diabetics and is a leading cause of end-stage kidney failure, contributing significantly to healthcare costs and cardiovascular morbidity [Sorvor et al., 2022, Adoba et al., 2025]. In Ghana, diabetes is a primary cause of kidney diseases, with a prevalence of 12.5% [Adoba et al., 2025]. Additionally, urinary tract infections (UTIs) are common opportunistic infections in diabetic patients globally, with an estimated 60 million cases among diabetics in 2019 [Kangume, 2024]. Diabetics are highly susceptible due to compromised immune systems and high urine glucose, which promotes bacterial growth [Kangume, 2024, Owusu and Afutu, 2022]. Antibiotic resistance among UTI pathogens further complicates management [Owusu and Afutu, 2022].

The quality of diabetes care is widely considered suboptimal, particularly in LMICs like Ghana, where studies indicate a high proportion of patients exceed recommended biochemical and clinical targets [Letsa, 2023]. Effective quality care, however, can substantially delay or prevent severe complications [Letsa, 2023]. The National Diabetes Quality Improvement Alliance (NDQIA) in the US established key diabetes care indicators in 2005, which are widely adapted for assessing healthcare performance. The six process indicators include glycated haemoglobin, LDL cholesterol, microalbuminuria, foot examination, dilated eye examination, and smoking status documentation [Letsa, 2023].

Despite increasing research on diabetes, a significant gap exists in the specific evaluation of diabetes care quality through validated indicators in LMICs, including Ghana [Letsa, 2023, Sorvor et al., 2022]. While microalbuminuria is an early marker for renal involvement in diabetes, its high prevalence in Ghana suggests an impending surge in complications [Eghan and Adjei-Poku, 2007]. A study in Kumasi found an alarmingly high prevalence of microalbuminuria at 43.1% among diabetic patients, with the median duration of diabetes before its development being 10 years [Eghan and Adjei-Poku, 2007]. Another study in the Kumasi metropolis, focusing on Type 2 Diabetes Mellitus (T2DM) patients, reported an even higher prevalence of microalbuminuria at 79.4% and macroalbuminuria at 13.1% [Sorvor et al., 2022]. However, a different study on urine biomarkers in diabetic patients in Ghana observed “mild albuminuria” in 10.4% of participants, noting this was significantly below the global prevalence for type 2 diabetics. Regarding overall kidney dysfunction, a cross-sectional study in the Obuasi East Municipality found the total prevalence to be 32.35%, comprising 23.5% with hyperfiltration (high GFR) and 8.82% with low GFR (<90 mL/min/1.73m²) [Adoba et al., 2025]. Nationally, diabetes mellitus is reported as a major cause of kidney diseases with a prevalence of 12.5% [Adoba et al., 2025]. Furthermore, a multicenter study revealed a prevalence of significant CKD (stages 3-5) at 44.3% among Ghanaian patients with hypertension and diabetes [Sorvor et al., 2022]. A systematic review and meta-analysis specifically for West Africa (including Ghana) reported a pooled prevalence of nephropathy of 11% among diabetes patients, which was the lowest regional prevalence across Africa [Sorvor et al., 2022].

There is limited data linking specific urine biomarkers to comorbidities and overall care quality, and a need for further studies on diagnostic efficacy of newer markers like serum free light chains [Sorvor et al., 2022]. Furthermore, detailed information on UTI prevalence, causative bacteria, and antimicrobial susceptibility patterns between diabetic patients and their non-diabetic caregivers in the same center has not been adequately established [Owusu and Afutu, 2022].

This study aims to address these critical research gaps by evaluating the quality of diabetes care at the Ghana Ports and Harbours Authority (GPHA) hospital using NDQIA-approved quality indicators [Letsa, 2023]. The general objective of this study was to through a medical audit, utilizing the quality indicators approved by the National Diabetes Quality Improvement Alliance (NDQIA) of the United States [Letsa, 2023].

## Methods

### Study Design and Setting

This study employed a cross-sectional design, primarily involving a retrospective medical audit of patient records at the Diabetes Clinic of the GPHA hospital in Ghana. The GPHA Diabetes Clinic is a weekly clinic at the GPHA hospital, Takoradi that sees approximately 30 to 40 clients weekly and is headed by a Senior medical doctor.

### Study Population and Sampling

The study population comprised clients whose urinalysis data were collected between 25 July 2022 and 30 November 2022, during which period Sysmex urinalysis analyzers UC 350 and UF 400 were procured and used. Participants were included if their diagnosis, retrieved from the hospital management system using their unique laboratory numbers, fell into one of the following categories: cyesis and diabetes only, diabetes and hypertension only, diabetes only, or cyesis diabetes hypertension only. This approach allowed for the linkage of specific laboratory findings to patient diagnoses.

### Data Collection Tools and Procedure

Data collection was conducted in two main parts

1. Urinalysis and Structural/Input Measures: Urinalysis Data: Demographic data (age and gender) as well as urinalysis data comprising urobilinogen, blood, bilirubin, ketones, protein, pH, nitrite, leukocyte, creatinine, albumin, protein creatinine ratio, albumin creatinine ratio, specific gravity, color, appearance, RTEC, conductivity and osmolality were retrieved from the Sysmex UC 350 urine analyser.
2. Clinical Process and Outcome Indicators: With approval from hospital authority and patient’s anonymised, a retrospective medical audit of the diagnosis of the selected patient records was retrieved by both a laboratory personnel and a nursing staff. Retrieved data was reviewed by a team of laboratory personnel to confirm data correctness and accuracy.

### Quality Control

Prior to data collection, research assistants received training on the study’s aims, data abstraction procedures, and ethical guidelines. The data abstraction tool was pre-tested on 50 subjects at the hospital to identify and rectify any gaps in the tool and procedure.

### Data Processing and Statistical Analysis

Collected data were first entered into Microsoft Excel 2021 and subsequently exported to Stata MP 17.0 for analysis. Descriptive statistics, such as means, standard deviations, frequencies, and proportions, were used to summarise both continuous and categorical variables. A univariate analysis of background characteristics was conducted and reported in numbers and percentages.

To assess the association between sociodemographic factors and the quality-of-care indicators (process and outcome indicators), Pearson’s Chi-square test was employed. A p-value of less than 0.05 was considered statistically significant.

### Ethical Considerations

Ethical approval for the study protocol was obtained from the management of the Ghana Ports and Harbours Authoriity hospital. For a retrospective study that does not directly involve participants, external ethical clearance was not sought. To ensure anonymity, no patient-identifiable information was used; sampled medical records were identified solely with unique laboratory identifiers.

## Results

### Demographics

A total of 125 participants, 52 (41.6%) males and 73 (58.4%) females were included in this study (Table 1). The age of study subjects ranged from 28 to 76 years with a mean of 54.92. Majority (55.2%) of the participants belonged to the 45-64 years category, 23.20% were above 64 years and the remaining 21.6% were in the 25-44 years group (Table 2). Further analysis showed that only 12% of the population were below 40 years. This information suggests that persons who are 40 years and above are at higher risk to diabetes in a similar population. Again, the study showed that 21.6% subjects below 45 years had diabetes. This is comparable to current global diabetes trends of 20-25% (WHO, 2024).

**Table 1:** Gender Distribution Across Population Categories.

| Population category | Male | Female | Total | Percent |
|---|---|---|---|---|
| Cyesis & diabetes | 0 | 5 | 5 | 4.00 |
| Cyesis, hypertension | 0 | 3 | 3 | 2.40 |
| Diabetes & hypertension | 14 | 7 | 21 | 16.80 |
| Diabetes only | 38 | 58 | 96 | 76.80 |
| Total | 52 | 73 | 125 | 100.00 |

**Table 2:** Age Distribution of Study Participants.

| Age range | Frequency | Percent | Cumulative Percent |
|---|---|---|---|
| 25–44 yrs | 27 | 21.60 | 21.60 |
| 45–64 yrs | 69 | 55.20 | 76.80 |
| 65 yrs and above | 29 | 23.20 | 100.00 |
| Total | 125 | 100.00 |  |

The varying diabetic categorizations represented in the study were diabetes only, diabetes and hypertension, diabetes and cyesis, and diabetes, hypertension and cyesis with corresponding numbers of 96 (76.8%), 21 (16.8%), 5 (4%), and 3 (2.4%) respectively (Table 1).

### Albuminuria, Glucosuria and Proteinuria Against the Varying Population

Although no significant correlation was established between albuminuria and diabetes population category (Pr=0.397), it is worth mentioning that overt albuminuria was absent for the study population and mild albuminuria was recorded for only 13 (10.4%) out of the 125 participants (Table 3). This is significantly below the 25-30% global prevalence of microalbuminuria among type-2 diabetics [Basi and Lewis, 2008]. Unfortunately, this study faced a challenge in acquiring the type of diabetes.

**Table 3:** Distribution of Albuminuria Across Population Categories.

| Albuminuria | Cyesis & diabetes | Cyesis, hypertension & diabetes | Diabetes & hypertension | Diabetes only | Total |
|---|---|---|---|---|---|
| Normal | 4 | 2 | 18 | 88 | 112 |
| Mild albuminuria | 1 | 1 | 3 | 8 | 13 |
| Total | 5 | 3 | 21 | 96 | 125 |

A significant level of correlation was established between the various diabetes population and urine glucose (Pr=0.001). About 82.4% (103) of the study subjects did not record glucose in their urine, 2.4% (3) had trace, and the remaining 15.2% (19) presented varying levels of glucosuria (Table 4). The non—availability of data on the type of medication or the level of complication is a limitation to comparing this finding to other similar studies.

**Table 4:** Distribution of Urine Glucose Levels Across Population Categories.

| Population category | Negative | 1+ | 2+ | 3+ | Trace | Total |
|---|---|---|---|---|---|---|
| Cyesis & diabetes | 4 | 0 | 0 | 1 | 0 | 5 |
| Cyesis, hypertension | 1 | 2 | 0 | 0 | 0 | 3 |
| Diabetes & hypertension | 17 | 1 | 0 | 2 | 1 | 21 |
| Diabetes only | 81 | 2 | 4 | 7 | 2 | 96 |
| Total | 103 | 5 | 4 | 10 | 3 | 125 |

No significant correlation was observed between the diabetic population categories and protein levels in urine (Pr=0.759). The study revealed that about 64.8% (81) of the study subjects did not record glucose in their urine, 10.4% (13) had trace, 9.6% (12) recorded a (+) increase in protein, 1.6% (2) recorded a (++) increase and the remaining 13.6% (17) presented a (+++) increase (Table 5).

**Table 5:**
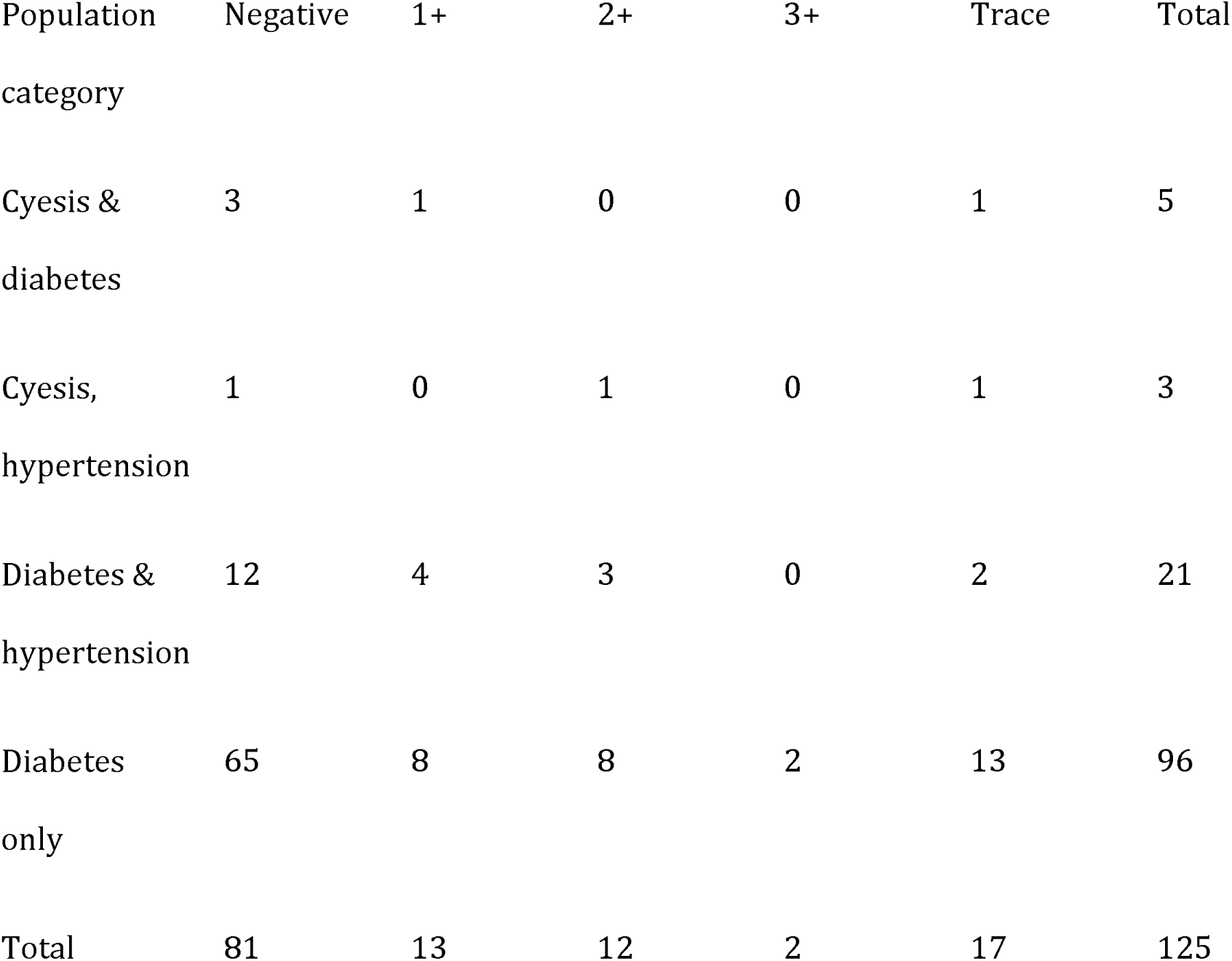
Distribution of Urine Protein Levels Across Population Categories.

| Population category | Negative | 1+ | 2+ | 3+ | Trace | Total |
|---|---|---|---|---|---|---|
| Cyesis & diabetes | 3 | 1 | 0 | 0 | 1 | 5 |
| Cyesis, hypertension | 1 | 0 | 1 | 0 | 1 | 3 |
| Diabetes & hypertension | 12 | 4 | 3 | 0 | 2 | 21 |
| Diabetes only | 65 | 8 | 8 | 2 | 13 | 96 |
| Total | 81 | 13 | 12 | 2 | 17 | 125 |

### Distribution of Renal Tubular Epithelial Cells (RTEC) Across Population Categories

Results on renal tubular epithelial cells (RTEC) was reported for only 26 participants of which 23 were normal (Table 6). Pathological results of 2.5 and above were reported for one cyesis and diabetes, and the remaining 2 was reported among diabetes and hypertension category.

**Table 6:** Distribution of Renal Tubular Epithelial Cells (RTEC) Across Population Categories.

| **Population category** | **Normal n (%)** | **≥2.5 n (%)** | **Total** |
|---|---|---|---|
| **Cyesis & diabetes** | 4 (17.39) | 1 (33.33) | 5 (19.23) |
| **Cyesis, hypertension** | 3 (13.04) | 0 (0.00) | 3 (11.54) |
| **Diabetes & hypertension** | 7 (30.43) | 2 (66.67) | 9 (34.62) |
| **Diabetes only** | 9 (39.13) | 0 (0.00) | 9 (34.62) |
| **Total** | **23 (100.00)** | **3 (100.00)** | **26 (100.00)** |

## Discussion

The primary findings of this study highlight the prevalence of key urine biomarkers—albuminuria, proteinuria, glucosuria, and Renal Tubular Epithelial Cells (RTEC)—among diabetic patients at the GPHA hospital and analyze their association with diabetic comorbidities. The demographic profile of the participants, showing a mean age of 54.92 years and a female majority (58.4%), is consistent with general observations in Ghana and the sub-region [Letsa, 2023]. For instance, a previous study in Kumasi reported a comparable mean age of 54.1 years [Eghan and Adjei-Poku, 2007]. The high female attendance (58.4%) aligns with documented trends where females are generally more proactive in seeking and reporting for regular health reviews compared to their male counterparts [Letsa, 2023].

### Interpretation of Renal Biomarkers

The evaluation of albuminuria revealed a low prevalence of early diabetic nephropathy markers in the studied population. Specifically, mild albuminuria (MA) was recorded in only 10.4% of participants, and overt albuminuria was completely absent (0.0%). This figure is significantly lower than the generally accepted global prevalence of microalbuminuria (25–30%) among type 2 diabetics. Furthermore, this finding contrasts markedly with other reports from Ghana, where a study in Kumasi found a microalbuminuria prevalence of 43.1% in diabetic patients, while another multicentre study reported a prevalence of significant CKD (stages 3–5) at 44.3% among Ghanaian patients with hypertension and diabetes [Eghan and Adjei-Poku, 2007, Sorvor et al., 2022]. The low prevalence observed in this study may be attributed to inherent limitations, including the small sample size and the inability to differentiate diabetes types (Type 1 vs. Type 2) or ascertain medication details, which limits comparison to external studies. Nonetheless, microalbuminuria is recognized as a crucial, early marker for renal involvement in diabetes and its monitoring is strongly recommended [Eghan and Adjei-Poku, 2007].

The study noted that overall kidney dysfunction (defined by GFR abnormalities) remains a significant concern in Ghana, with a cross-sectional study in the Obuasi East Municipality reporting a total prevalence of 32.35%, comprising 23.5% hyperfiltration and 8.82% low GFR (<90 mL/min/1.73m²) [Adoba et al., 2025]. Regionally, a systematic review noted that the pooled prevalence of nephropathy in West Africa (including Ghana) was 11%, the lowest among African regions studied [Sorvor et al., 2022]. The detection of pathological Renal Tubular Epithelial Cells (RTEC) in a few individuals (3 out of 26 tested), particularly those with concurrent diabetes and hypertension, suggests that structural kidney damage indicators appear primarily in patients with established comorbidities.

### Glucosuria and Comorbidity Association

A notable outcome was the **significant correlation found between the various diabetic population categories and urine glucose** (Pr=0.001). While a majority of subjects (82.4%) recorded no urine glucose, 15.2% presented varying levels of glucosuria. This association indicates that the severity or complexity of the diabetic condition, reflected by comorbidity (e.g., hypertension, cyesis), influences glycemic control as measured by urinary glucose excretion. The presence of glucose in the urine promotes bacterial growth, significantly increasing the diabetic patient’s susceptibility to Urinary Tract Infections (UTIs) due to the compromised immune system associated with DM [ Kangume, 2024]. The association between UTIs and female gender, disease comorbidities, and late diabetic disease has been reported elsewhere [Kangume, 2024]. Furthermore, similarities in UTI prevalence and bacterial uropathogens found in diabetic patients and their non-diabetic caregivers suggest the circulation of antibiotic-resistant strains within the healthcare environment [Owusu and Afutu, 2022].

### Limitations and Future Directions

The study was limited by its cross-sectional design, the small sample size, and the challenge in documenting the specific type of diabetes, medication, and the level of complication, which restricts a comprehensive comparison against NDQIA quality outcome targets. Nevertheless, the focus on specific urinary biomarkers provides unique baseline data for the GPHA clinic. Future efforts should emphasize addressing the research gap regarding the quality of diabetes care specifically evaluated through comprehensive care indicators [ Letsa, 2023]. The continued evaluation of diagnostic efficiency for newer markers like serum free light chains (which showed promise but required further elucidation in other Ghanaian studies) remains important for early detection of DN [Sorvor et al., 2022].

### Conclusion

Diabetes mellitus continues to impose a significant health burden globally, requiring optimal quality of care to mitigate its associated complications and mortality [Letsa, 2023]. This study at the GPHA hospital found that the prevalence of mild albuminuria (10.4%) was remarkably low compared to established international and many national benchmarks. Conversely, a **significant correlation was established between the complexity of the diabetic population (comorbidities) and urine glucose levels** (Pr=0.001). Given that kidney involvement in diabetes is a major cause of subsequent morbidity and mortality, continued adherence to strict monitoring guidelines is imperative [Eghan and Adjei-Poku, 2007]. Identifying these specific associations between diabetic categories and glucosuria provides localized data crucial for developing targeted management and preventive strategies at the GPHA hospital and similar clinics. Further research evaluating the full spectrum of NDQIA quality indicators across diabetic clinics nationally is necessary to address underlying systemic shortcomings in diabetes care [Letsa, 2023].

## Data Availability

All data produced in the present study are available upon reasonable request to the authors

## Acknowledgements

The authors are grateful to the management of GPHA Hospital, Takoradi, for granting ethical and administrative approval to undertake this study. We also appreciate the support of the hospital staff who facilitated access to the required data.

## Abbreviations

DM, diabetes mellitus; DN, diabetic nephropathy; GPHA, Ghana Ports and Harbours Authority; LMICs, low- and middle-income countries; NDQIA, National Diabetes Quality Improvement Alliance; RTEC, renal tubular epithelial cells; T2DM, type 2 diabetes mellitus; UTIs, urinary tract infections.

